# The plasma metabolome of long COVID-19 patients two years after infection

**DOI:** 10.1101/2023.05.03.23289456

**Authors:** Yamilé López-Hernández, Joel Monárrez Aquino, David Alejandro García López, Jiamin Zheng, Juan Carlos Borrego, Claudia Torres-Calzada, José Pedro Elizalde-Díaz, Rupasri Mandal, Mark Berjanskii, Eduardo Martínez-Martínez, Jesús Adrián López, David S. Wishart

## Abstract

**Background:** One of the major challenges currently faced by global health systems is the prolonged COVID-19 syndrome (also known as “long COVID”) which has emerged as a consequence of the SARS-CoV-2 epidemic. The World Health Organization (WHO) recognized long COVID as a distinct clinical entity in 2021. It is estimated that at least 30% of patients who have had COVID-19 will develop long COVID. This has put a tremendous strain on still-overstretched healthcare systems around the world.

**Methods:** In this study, our goal was to assess the plasma metabolome in a total of 108 samples collected from healthy controls, COVID-19 patients, and long COVID patients recruited in Mexico between 2020 and 2022. A targeted metabolomics approach using a combination of LC-MS/MS and FIA MS/MS was performed to quantify 108 metabolites. IL-17 and leptin concentrations were measured in long COVID patients by immunoenzymatic assay.

**Results:** The comparison of paired COVID-19/post-COVID-19 samples revealed 53 metabolites that were statistically different (FDR < 0.05). Compared to controls, 29 metabolites remained dysregulated even after two years. Notably, glucose, kynurenine, and certain acylcarnitines continued to exhibit altered concentrations similar to the COVID-19 phase, while sphingomyelins and long saturated and monounsaturated LysoPCs, phenylalanine, butyric acid, and propionic acid levels normalized. Post-COVID-19 patients displayed a heterogeneous metabolic profile, with some showing no symptoms while others exhibiting a variable number of symptoms. Lactic acid, lactate/pyruvate ratio, ornithine/citrulline ratio, sarcosine, and arginine were identified as the most relevant metabolites for distinguishing patients with more complicated long COVID evolution. Additionally, IL-17 levels were significantly increased in these patients.

**Conclusions:** Mitochondrial dysfunction, redox state imbalance, impaired energy metabolism, and chronic immune dysregulation are likely to be the main hallmarks of long COVID even two years after acute COVID-19 infection.

## Background

Historically, highly pathogenic beta-coronaviruses have been associated with severe respiratory diseases. According to the WHO, the severe acute respiratory syndrome coronavirus (SARS-CoV), and the Middle East respiratory syndrome coronavirus (MERS-CoV) were responsible for epidemics in 2002-2003 and 2015, respectively. During the SARS-CoV epidemic, the virus was reported in 29 countries with 8,437 cases and 813 fatalities [1]. On the other hand, MERS-CoV was reported in 27 countries with 2,519 laboratory-confirmed cases between 2012 and 2020, resulting in 866 deaths [2]. In 2019, exactly 100 years after the last pandemic caused by an H1N1 influenza A virus (the Spanish flu), a new pandemic affected almost every country around the world. As of February 26, 2023, over 758 million confirmed cases of SARS-CoV-2 and over 6.8 million deaths have been reported globally. To date, around 653 million patients have recovered [3]. However, as early as spring 2020, people began describing their experiences of not fully recovering from SARS-CoV-2 infection [4]. This extended version of the disease has been called “long COVID”. Interestingly, the term long COVID is a patient-created term promoted in Twitter by Elsa Perego, an archeologist at University College London.

It has been widely described that some viruses lead to persistent physiological alterations even a decade after infection. The term “post-viral syndrome” has been in use for over a century [5]. Chronic symptoms such as fatigue, joint pain, and cardiovascular problems have been reported after recovery from other infections such as the West Nile, Polio, Dengue, Zika, seasonal flu, Epstein-Barr, Ebola, MERS, and SARS [6, 7]. However, none of these viruses have affected so many people in the same time window as SARS-CoV-2, which offers the scientific community a unique opportunity to understand the etiology of post-viral syndromes such as long COVID.

Long COVID (also known as post-COVID-19 syndrome or post-acute sequelae of COVID-19 (PACS)) is a condition characterized by long-term or persistent health problems appearing after the initial recovery from COVID-19 infection. The WHO has described long COVID as a condition “that occurs in individuals with a previous history of probable or confirmed SARS-CoV-2 infection, usually three months after the onset, with symptoms lasting at least two months that cannot be explained by an alternative diagnosis” [8]. It is estimated that 30-60% of recovered patients, even after a mild disease, will experience long COVID or symptoms persistence with varying durations [9]. Based on a conservative estimated incidence, at least 65 million individuals worldwide could be experiencing long COVID [3].

Similar to COVID-19, long COVID affects multiple organ systems, including the respiratory, cardiovascular, nervous, and gastrointestinal systems. More than 50 symptoms have been reported associated with long COVID [10]. Observational studies have reported that some symptomatic conditions are resolved within three months of hospitalization in 50% of patients [11], but the rate of full recovery drops to 35% between three and six months after hospitalization, and to 15% between six to nine months. Importantly, a high proportion of that population has residual lung tissue injury, with detectable radiological abnormalities on chest computed tomography (CT) scans [12]. Fatigue, loss of concentration, headaches, shortness of breath, anosmia, muscle weakness, joint pain are some of the symptoms most reported. Therefore, more than a homogeneous entity, long COVID could be considered as a spectrum of disorders, that affects individuals with complications directly linked to the virus (long-term residual damage in the lungs, brain, or heart), and individuals manifesting systemic unspecific signs/symptoms (fatigue, headache, and arthromyalgias) [13]. The increasing number of patients with long COVID poses a challenge for public health systems around the world, but, currently, there are no guidelines for accurately diagnosing patients with long COVID and classification is still underestimated and subjective.

In the present work we used quantitative targeted metabolomics to evaluate the metabolic reversion of patients with persistent sequelae due to confirmed SARS-CoV-2 infection. Comparison with negative controls allowed us to identify those metabolites persistently dysregulated after two years of the initial infection. Number, type of symptoms as well as metabolic signatures were different in patients experiencing long COVID. Besides, IL-17 level was increased in patients with the worst disease evolution. To the best of our knowledge, this is the first targeted metabolomics study of long COVID patients conducted beyond twenty months post-infection.

## Methods

### Patient recruitment

For the aims of this study, COVID-19 patient survivors (with confirmed diagnostic based on a positive PCR for SARS-CoV-2) who developed a mild, severe, or critical disease, and were admitted (or hospitalized) in the Instituto Mexicano de Seguridad Social (Zacatecas city, Mexico) and Christus Muguerza del Parque Hospital (Chihuahua city, Mexico) between March and November 2020, were recruited. Participants were contacted for a face-to face interview. They were invited to respond to a questionnaire and to donate a blood sample. Plasma was isolated from the donated blood. COVID-19 patients from the Instituto Mexicano de Seguridad Social were recruited from an initial set of 124 COVID-19 patients enrolled in a previous research study [14]. Of these, 44 (35.6%) passed away during hospitalization and in the following months after hospital discharge. From 80 survivors, it was possible to contact 36 (by their Social Security Number or personal/relative phone number kept in hospital records), and 15 agreed to participate. For these 15 patients, paired plasma samples from the first diagnosis of the acute disease (COVID-19 group) and post-COVID phase were available.

Additionally, from a cohort of patients that were hospitalized in 2020 in Christus Muguerza del Parque Hospital, 33 were randomly selected by age stratification. For those patients, a basal blood sample was not available; however, all clinical information and chest computed tomography (CT) scans were recorded in the hospital archive.

For the neuropsychological assessment, the validated Hamilton Anxiety Rating Scale (HAM-A) [15] was used. For depression assessment, the Hamilton Depression scale (HAM-D) was used [16]. The Montreal Cognitive Assessment (MoCA) was employed for cognitive impairment [17]. For dyspnea assessment, the modified Medical Research Council (mMRC) dyspnea scale was implemented [18]. Basic blood biochemical markers were performed (i.e., hemoglobin, platelets, leukocytes, lymphocytes, and creatinine) for all enrolled patients.

To assess for differences in the severity of long COVID patients, our own classification was made (arbitrarily) considering the frequency of concomitant symptoms. Recovered patients were classified as those who did not report persistent symptoms. Long COVID was considered if patients reported at least one persistent neurologic, psychiatric, gastrointestinal, cardiologic, respiratory, or systemic symptom. The Class A long COVID patients were those reporting less than five persistent symptoms (17 patients), while class B long COVID patients were those reporting five or more persistent symptoms (13 patients). As negative controls and an indicator of normal population, stored plasma samples from 37 individuals who tested negative for SARS-CoV-2 in 2020 were used.

This study was conducted in accordance with the Declaration of Helsinki (1976). It was also revised and approved by the Research and Ethics Committees of the Instituto Mexicano de Seguridad Social, with the registration number R-2022-3301-038, and Christus Muguerza del Parque Hospital (HCMP-CEI-15042020-3, and HCMP-CEI-28022022-A01). Informed consent was obtained from all participants. All patients included in this study were informed in writing regarding the collection of their samples for research aims and were given the right to refuse participation.

### Metabolomics analysis

A combination of direct injection mass spectrometry with a reverse-phase LC-MS/MS custom assay was used, as previously described [14]. Briefly, metabolites were measured using a locally developed LC–MS/MS metabolomics assay called The Metabolomics Innovation Centre (TMIC) Prime (TMIC PRIME®) Assay. This assay provides quantitative results for up to 143 endogenous metabolites, including biogenic amines, amino acids, organic acids, lipids, and lipid-like compounds.

The method combines the derivatization and extraction of analytes, and the selective mass-spectrometric detection using multiple reaction monitoring (MRM) pairs. Isotope-labeled internal standards and other internal standards were used for metabolite quantification. The custom assay uses a 96 deep-well plate with a filter plate attached via sealing tape, and reagents and solvents used to prepare the plate assay. The first 14 wells of the 96-well plate were used for calibration and quality control with one double blank, three zero samples, seven calibration standards and three quality control samples. To measure all metabolites except organic acids, samples were first thawed on ice and were vortexed. 10 µL of each sample was loaded onto the center of the filter on the upper 96-well plate and dried under a stream of nitrogen. Subsequently, phenyl-isothiocyanate (PITC) was added for derivatization. After incubation, the filter spots were dried again using an evaporator. Extraction of the metabolites was then achieved by adding 300 µL of extraction solvent. The extracts were obtained by centrifugation into the lower 96-deep well plate, followed by a dilution step with the mass spectrometry running solvent.

For organic acid analysis, 150 µL of ice-cold methanol and 10 µL of isotope-labeled internal standard mixture was added to 50 µL of each plasma sample for overnight protein precipitation. Each sample was then centrifuged at 13000 x g for 15 min. 50 µL of supernatant was loaded into the center of wells of a 96-deep well plate, followed by the addition of ^13^C labeled 3-nitrophenylhydrazine (3-NPH) as an isotopic labeling reagent (for quantification). After incubation for 2h, butylated hydroxytoluene (as a stabilizer) and water were added before LC-MS injection.

Mass spectrometric analysis for the PITC-derivatized and 3-NPH-derivatized samples was performed on an ABSciex 4000 Qtrap® tandem mass spectrometry instrument (Applied Biosystems/MDS Analytical Technologies, Foster City, CA) equipped with an Agilent 1260 series UHPLC system. Organic acids, biogenic amines, amino acids, and amino acid derivatives were detected and quantified via LC-MS, while lipids, acylcarnitines, and glucose were detected and quantified via a direct injection (DI) method.

Analyst 1.6.2 and MultiQuant 3.0.3 was used for quantitative analysis. An individual seven-point calibration curve was generated to quantify organic acids, amino acids, biogenic amines, and derivatives. Ratios for each analyte’s signal intensity to its corresponding isotope-labelled internal standard were plotted against the specific known concentrations using quadratic regression with a 1/x^2^ weighting. For lipids, acylcarnitines, and glucose, a single point calibration of a representative analyte was built using the same group of compounds that share the same core structure assuming a linear regression through zero.

### Plasma IL-17 and leptin determinations

ELISA kits were used for the quantification of IL-17 (Catalog Number RAB0262, Sigma-Aldrich, St. Louis, MO, USA) and leptin (catalog number ab108879, Abcam, Cambridge, UK), following manufacturer’s instructions. Briefly, standard solutions (or plasma samples), were added to each type of pre-coated 96-well plate and incubated overnight at 4°C. The plates were then incubated with the corresponding detection antibodies (100 μL/well) for 1 h at room temperature. Streptavidin solution (100 μL) was then added to each well and the plates were incubated for 45 min. After the antibody-HPR incubation, TMB one-step substrate reagent (100 μL) was added to the wells and the plates were incubated for another 30 min before the addition of a stop solution (50 μL/well). Absorbance values (at 450 nm) were used for the calculation of the protein concentrations (pg/mL) by comparing the absorbance to an appropriate standard curve.

### Statistical analysis

To describe baseline characteristics of negative controls (non-COVID-19), COVID-19 or post-COVID-19 patients, medians with interquartile ranges (IQRs) or means [with standard deviations (s.d.)] and frequencies (%) were used for continuous and categorical data, respectively. Normality was assessed using the D’Agostino-Pearson normality test. Student’s t-test or Mann-Whitney tests were used for continuous data. For categorical variables (e.g., sex, smoking, symptoms, and comorbidities) Pearson Chi^2^ tests or Fisher’s exact tests were used. All p-values less than 0.05 (p<0.05) were considered statistically significant. Analyses were conducted using SPSS (IBM, version 24).

Metabolite analysis was performed with MetaboAnalyst 5.0 [19]. Those metabolites with more than_20% of missing values were removed from further analysis. For the remaining metabolites, values below the limit of detection (LOD) were imputed using 1/5 of the minimum positive value of each variable. The data were then subject to median normalization, log-transformed and Pareto-scaled to generate appropriate Gaussian metabolite concentration distributions. Differences in mean metabolic values between controls, COVID-19, post-COVID-19, and long COVID patients were assessed using a parametric t-test or one-way ANOVA [adjusted p-value (FDR) cut-off = 0.05]. For the paired study, t-test, and volcano plots of log-transformed p-values were generated to address significant metabolites. Principal component analysis (PCA) and two-dimensional partial least squares discriminant analysis (2-D PLS-DA) scores plots were used to compare plasma metabolite data across and between study groups; 2000-fold permutation tests were used to assess statistical significance and minimize the possibility that the observed separation of the PLS-DA clusters was due to chance. Differentiated metabolites were identified by a variable importance in projection (VIP) using a score cutoff of >1.5. Heat maps of the top 50 significant metabolites (via t-test or ANOVA) were created via MetaboAnalyst.

Pathway analysis was done using Metabolite Set Enrichment Analysis (MSEA) and Metabolomic Pathway Analysis (MetPA) modules as found in MetaboAnalyst 5.0 [19]. The Homo sapiens pathway library was used for pathway analysis. The global test was used for the selected pathway enrichment analysis method, whereas the node importance measure for topological analysis was used to assess the relative betweenness centrality.

The metabolites with the highest VIP scores and LASSO frequencies were used to create metabolite panels for predicting long COVID using multivariate logistic regression.

Additionally, models were adjusted for relevant potential confounders such as sex, age, relevant comorbidities (i.e., DM-II, HTN, and obesity), so that only statistically significant variables (p_<_0.05) remained in the final models. Logistic regression analysis was performed with the Pareto-scaled data. K-fold cross-validation (CV) was used to ensure that the logistic regression models were robust. To determine the performance of each generated model, the area under the receiver operating characteristics curve (AUROC or AUC) was calculated, as was sensitivity and specificity.

## Results

### Demographic, clinical data and symptoms description

**Table 1** shows baseline characteristics of patients enrolled in the study. Age was statistically different between negative (healthy) controls and COVID-19 patients. However, differences were not found in the self-reported comorbidities. Six patients (12.5%) developed mild disease; 37 patients developed (77%) moderate/severe disease while five patients (10.4%) developed critical disease. Six patients (12.5%) were reinfected during 2021 and 2022. All patients were fully vaccinated during the period of 2021-2022. The questionnaire answered by the patients revealed the most persistent symptoms which were grouped into five broad categories: systemic, neurologic, psychiatric, cardiologic, and respiratory. The most predominant symptoms were loss of memory (73.3%), sleep disorders, arthralgia, fatigue, exercise intolerance, myalgia (66.7%), and anxiety (60.0%).

**Table 1:** Baseline characteristics of participants in the study.

| Variable | Controls<br>(N=38) | COVID-19<br>(N=15) | post-COVID-19<br>(N=48) | long COVID<br>(N=29) | p-value |
| --- | --- | --- | --- | --- | --- |
| Age, median (Q1- | 40.5 (37- | 51 (44.3-59.3) | 51.5 (43.5-60.8) | 50.5 (44.3-62) | 0.0079 *<sup>a, b, c</sup> |
| Q3) | 53.3) |  |  |  |  |
| Male gender, n (%) | 17 (44.7) | 12 (54.5) | 28 (58.3) | 9 (34.6) | 0.2226 |
| Smoking, n (%) | 4 (10.5) | 3 (13.6) | 7 (14.6) | 3 (11.5) | 0.7231 |
| **Comorbidities (self-reported), n (%)** |  |  |  |  |  |
| Diabetes | 3 (7.9) | 3 (13.6) | 7 (14.6) | 3 (11.5) | 0.7833 |
| Hypertension | 9 (23.7) | 6 (27.3) | 20 (41.7) | 9 (34.6) | 0.1035 |
| Obesity | 3 (7.9) | 0 (0) | 5 (10.4) | 2 (7.7) | 0.3823 |
| **Laboratory data, median (Q1-Q3)** |  |  |  |  |  |
| Hemoglobin (g/dL) | 15.3 (14.4-16.2) | 14.4 (12.6-16.2) | 15.4 (14.5-16.1) | 15.5 (14.68-16.23) | 0.7833 |
| Platelets (thousands/ mL) | 276 (236.5-325.5) | 177 (68.2-281.8) | 242.5 (219.8-290.5) | 242.3 +/- 69.7 | 0.1035 |
| Leukocytes ($\times 10^3$) | 7.1 (6.0-8.3) | 6.7 (4.4-11.1) | 7.0 (6.2-7.8) | 7.3 (6.5-7.7) | 0.3823 |
| Lymphocyte counts (%) | 31.8 (25.5-36.4) | 12.1 (4.9-22.1) | 33.8 (29-38.9) | 34 +/-7.1 | <0.0001 *a, d, e |
| Creatinine (mg/dL) | 0.9 (0.7-1.1) | 0.7 (0.6-0.9) | 0.8 (0.7-1.0) | 0.8 (0.7-0.9) | 0.2452 |

a: negative controls vs. COVID-19; b: COVID-19 vs. post-COVID-19; c: negative controls vs. post-COVID-19; d: COVID-19 vs. Post-COVID-19; e: COVID-19 vs. long COVID; f: Post-COVID-19 vs. long COVID

### Paired analysis: COVID-19/post-COVID-19 phases

When paired samples (COVID-19/post-COVID-19) from 15 patients were compared metabolically, 53 plasma metabolites were found to be significantly different (FDR < 0.05). **Supplementary Table 1** shows the results of the t-test (autoscaling normalization). The volcano plot **(Figure 1A)** shows that 13 metabolites were significantly upregulated in the post-COVID-19 phase, while 32 were downregulated with a fold change (FC) threshold > 1.3 (FDR <0.05). Heatmap analysis **(Figure 1B)** shows a clustering of patients corresponding to their COVID-19 and post-COVID phases, revealing that lysoPCs (except LysoPC 18:2) and SMs were downregulated in the post-COVID phase. Multivariate analysis (via PLS-DA) demonstrated a clear separation between both COVID phases (accuracy: 0.97, R^2^: 0.94, Q^2^: 0.77) **(Figure 1C)**. The VIP plot **(Figure 1D)** shows that phenylalanine, taurine, and spermidine had lower plasma concentrations in the post-COVID-19 phase, while the glutamine/glutamate ratio was increased in the post-COVID-19 phase.

**Figure 1:**
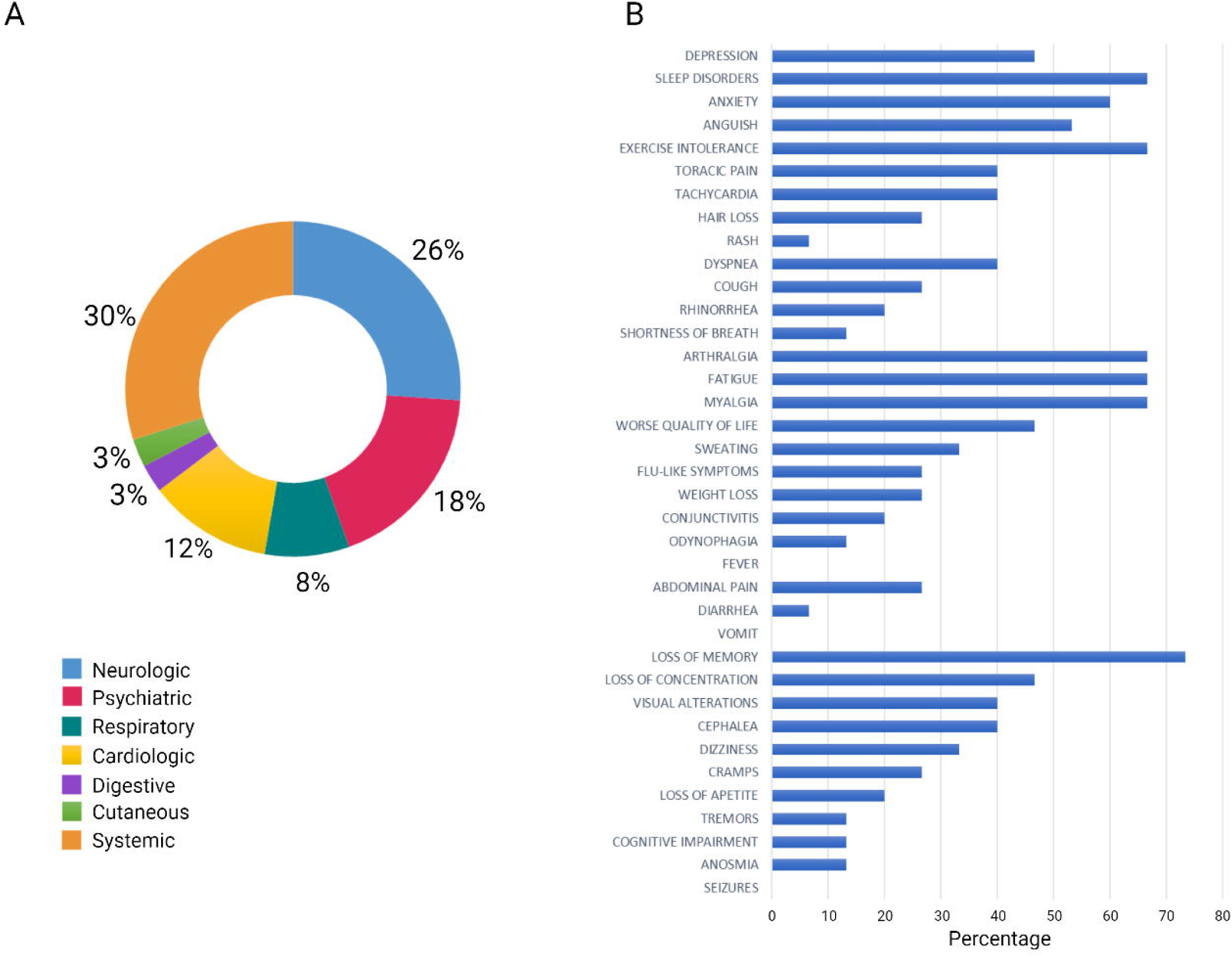
Most common symptoms remaining after 24 months in 48 post-COVID-19 patients. A) Distribution of symptoms according to organ systems. B) Results obtained via in-person questionnaire.

### Post-COVID-19 patients compared with controls

In order to know if the altered metabolites (and those that remained unsignificant) were dysregulated with respect to normal values, a group of negative SARS-CoV-2 controls (i.e., healthy controls) collected from 2020 was added to the analysis. The heatmap **(Figure 2A)** shows the metabolites with significant differences across the three study groups (one way ANOVA). Volcano plots **(Figure 2B)** were used to identify the dysregulated metabolites in the post-COVID-19 patients with respect to the healthy controls. Two-sample t-tests & Wilcoxon rank-sum tests shows that in comparison with the healthy controls, 29 metabolites were still dysregulated in post-COVID patients **(Supplementary Table 2)**. C18:1(adjusted p = 4 x 10^-3^), C18:2 (adjusted p = 6.1 x 10^-6^), C10:2 (adjusted p = 7 x 10^-3^), C10:1(adjusted p = 3 x 10^-2^), carnitine (adjusted p = 4 x 10^-2^) as well as glutamine (adjusted p = 4.2 x 10^-5^), choline (adjusted p = 5.7 x 10^-5^), glucose (adjusted p = 4.2 x 10^-5^), kynurenine (adjusted p = 7 x 10^-4^), pyruvic acid (adjusted p = 1.0 x 10^-3^), kynurenine/tryptophan ratio (adjusted p = 6.0 x 10^-03^), threonine (adjusted p = 7 x 10^-3^), putrescine (adjusted p = 0.01), ornithine (adjusted p = 0.04), PC ae 36:0 (adjusted p = 0.03), and PC aa 40:2 (adjusted p = 0.02) were found in higher concentrations in post-COVID-19 patients relative to the healthy controls.

**Figure 2:**
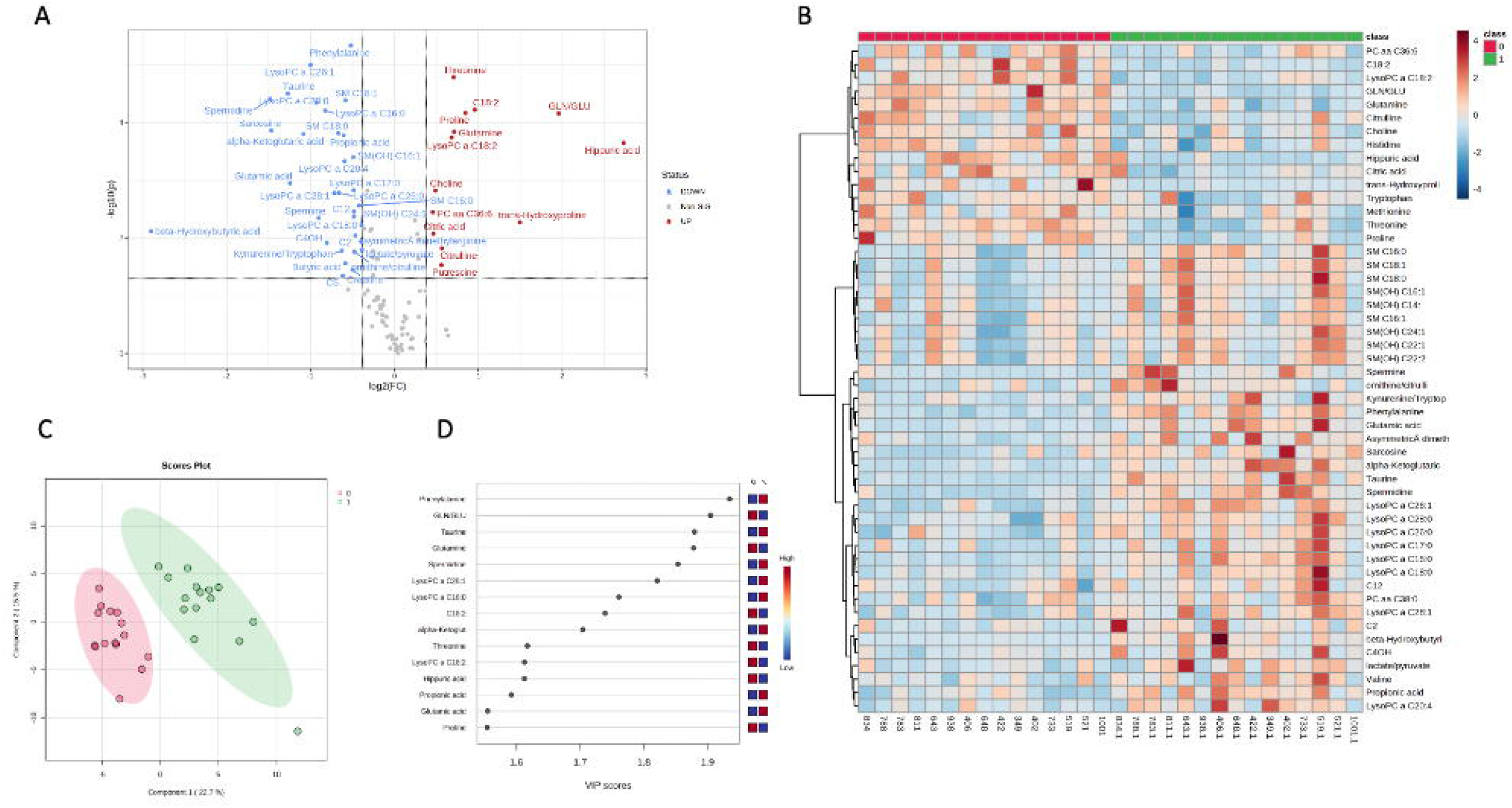
Multivariate analysis. A) The volcano plot of the plasma metabolomics between the COVID-19 phase and the post-COVID-19 phase (red represents the up-regulated metabolites compared with COVID-19 phase, green represents the downregulated metabolites compared with COVID-19 phase, and gray represents the metabolites with no difference between both groups. Fold change threshold_=_1.3 and p-value = 0.05 (FDR adjusted). B) Representative heatmap of top 50 significant metabolites (t-test) in the comparison of COVID-19 and post-COVID-19 phases (red: post-COVID-19 phase; green: COVID-19 phase). C) Score scatter plot based on the two-dimensional PLS-DA (red: post-COVID-19 phase; green: COVID-19 phase) D) Rank of the different metabolites (the top 15) identified by the PLS-DA according to the VIP score on the x-axis. The most discriminating metabolites are shown in descending order of their coefficient scores. The color boxes indicate whether metabolite concentration is increased (red) or decreased (blue).

In addition, lysoPC 14:0 (adjusted p = 0.01), lysoPC 16:0 (adjusted p = 1.4 x 10^-11^), lysoPC 16:1(adjusted p = 1.0 x 10^-3^), lysoPC 18:0 (adjusted p = 3.0 x 10^-6^), LysoPC 17:0 (adjusted p = 1.0 x 10^-4^), LysoPC 20:4 (adjusted p = 2.0 x 10^-3^), SM(OH)22:1(adjusted p = 0.04), PC aa 32:2 (adjusted p = 0.01), sarcosine (adjusted p = 1.04 x 10^-5^), taurine (adjusted p = 4.2 x 10^-5^), and glutamic acid (adjusted p = 1.0 x 10^-3^) were found downregulated in post-COVID-19 patients relative to the healthy controls. Likewise, the glutamine/glutamate ratio (adjusted p = 2.7 x 10^-6^) was increased in post-COVID-19 patients, while the lactate/pyruvate ratio (adjusted p = 0.04) was found to be decreased.

Glucose, kynurenine, C10:2, C18:1, C10:1, lysoPC14:0, lysoPC16:1, PC ae 36:0, and PC aa 32:2, PC aa 40:2, were found to be in similar concentration levels as for those in the COVID-19 phase.

Several other metabolites previously related with severity in COVID-19 tend towards normal or healthy levels (kynurenine/tryptophan ratio, C18:2, glutamic acid, glutamine, spermidine) in the post-COVID-19 group. Of note, a group of sphingomyelins (SM(OH)14:0, SM16:0, SM(18:0), SM(OH)16:1, SM(OH)24:1, SM(18:1), SM(16:1)) were found to be normalized, as well as lysoPC 26:0, lysoPC 26:1, lysoPC 28:1, and lysoPC 28:0, phenylalanine, butyric acid, and propionic acid.

The multivariate analysis (PLS-DA) showed a clear separation between both classes (accuracy: 1; R^2^: 0.98; Q^2^:0.89) **(Figure 2C)**. The VIP score plot **(Figure 2D)** shows that the most important variables that can be used to differentiate negative controls from post-COVID-19 patients are glutamine/glutamate ratio, sarcosine, C18:2, taurine and LysoPC16:0.

A logistic regression model was built using both symptoms and plasma metabolites to distinguish the development of long COVID among COVID-19 patients. The AUC, sensitivity, and specificity values with 95% CI for this model are shown in Supplementary Table 3. A logistic regression model had the following equation: logit(P) = log(P / (1 - P)) = 1.13 - 1.724 myalgia - 2.763 ornithine/citrulline + 3.076 lactate/pyruvate, where the numeric value of each named metabolite in the equation is the concentration after auto-scaling.

### Investigating post-COVID-19 patients

Differences were found in patients from the post-COVID-19 group, both in the frequency of symptoms reported and in the plasma levels of some metabolites such as lactic acid, with a bimodal distribution across the group **(Supplementary Figure 1)**. Therefore, these patients were subclassified according to our own scale as a surrogate for disease severity. 18 patients did not report any symptoms (recovered). 17 patients reported less than five persistent symptoms (class A long COVID), while 13 reported more than five symptoms (class B long COVID).

**Figure 4** shows the box and whisker plots based on one way ANOVA for class A, class B, and fully recovered patients. The lactate/pyruvate ratio (adjusted p value = 5.8 x 10^-7^), lactate (adjusted p value = 4.8 x 10^-6^), arginine (1.8 x 10^-3^), ornithine/citrulline ratio (adjusted p value 5. x 10^-3^), and sarcosine (adjusted p value = 0.02) were the variables best able to differentiate long COVID patients with more than five symptoms from patients with less than five symptoms. Arginine and sarcosine negatively correlated with the number of symptoms.

**Figure 3:**
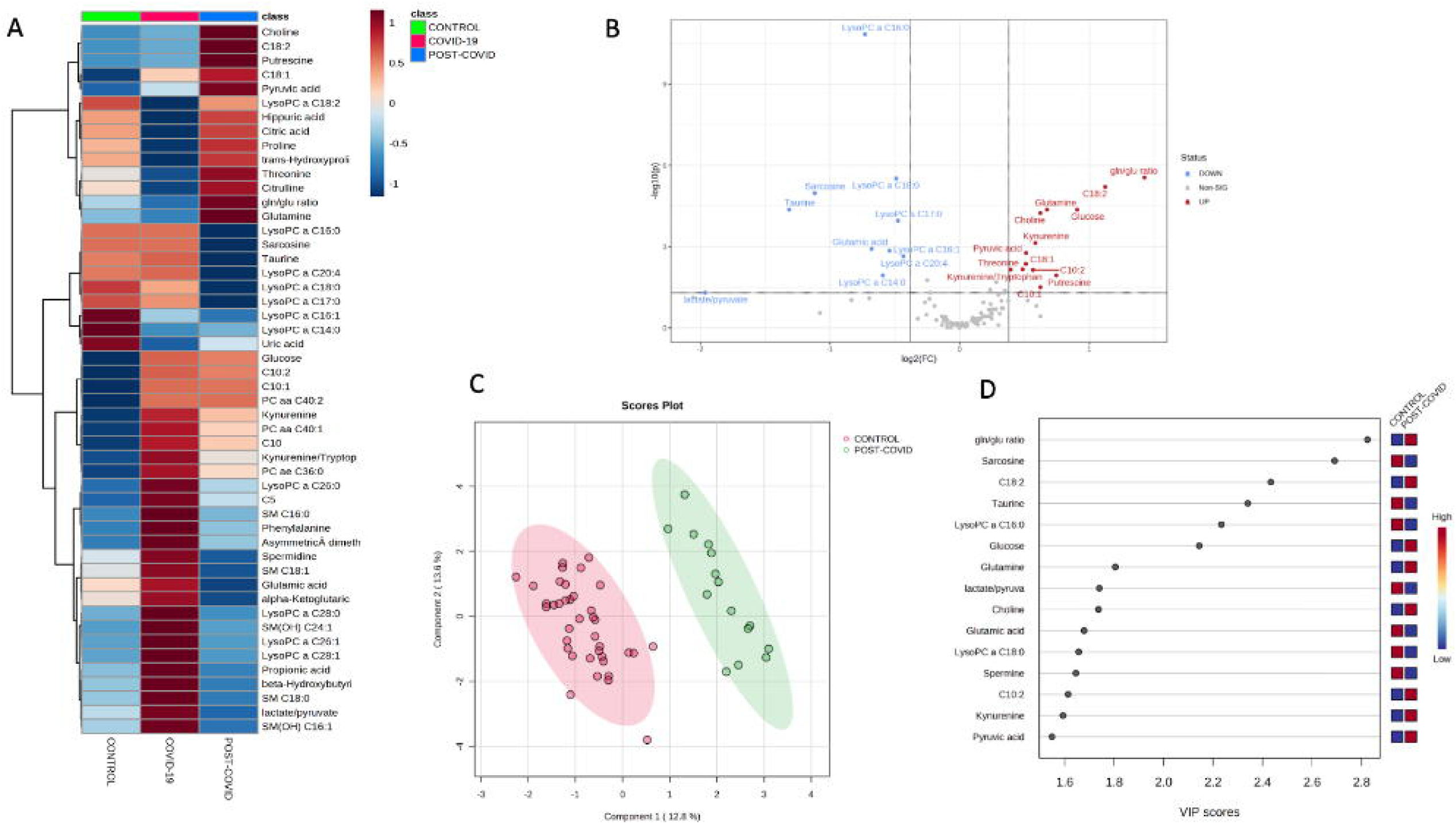
Multivariate analysis. A) Representative heatmap of top 50 significant metabolites (ANOVA) in the comparison of controls, COVID-19 and post-COVID-19 patients. B) The volcano plot of the plasma metabolomics between controls and post-COVID-19 patients (red represents the up-regulated metabolites compared with controls, green represents the down-regulated metabolites compared with controls, and gray represents the metabolites with no difference between both groups. Fold change threshold_=_1.3 and p-value =0.05 (FDR adjusted). C) Score scatter plot based on the two-dimensional PLS-DA (red: controls; green: post-COVID-19 patients) D) Rank of the different metabolites (the top 15) identified by the PLS-DA according to the VIP score on the x-axis. The most discriminating metabolites are shown in descending order of their coefficient scores. The color boxes indicate whether metabolite concentration is increased (red) or decreased (blue).

**Figure 4:**
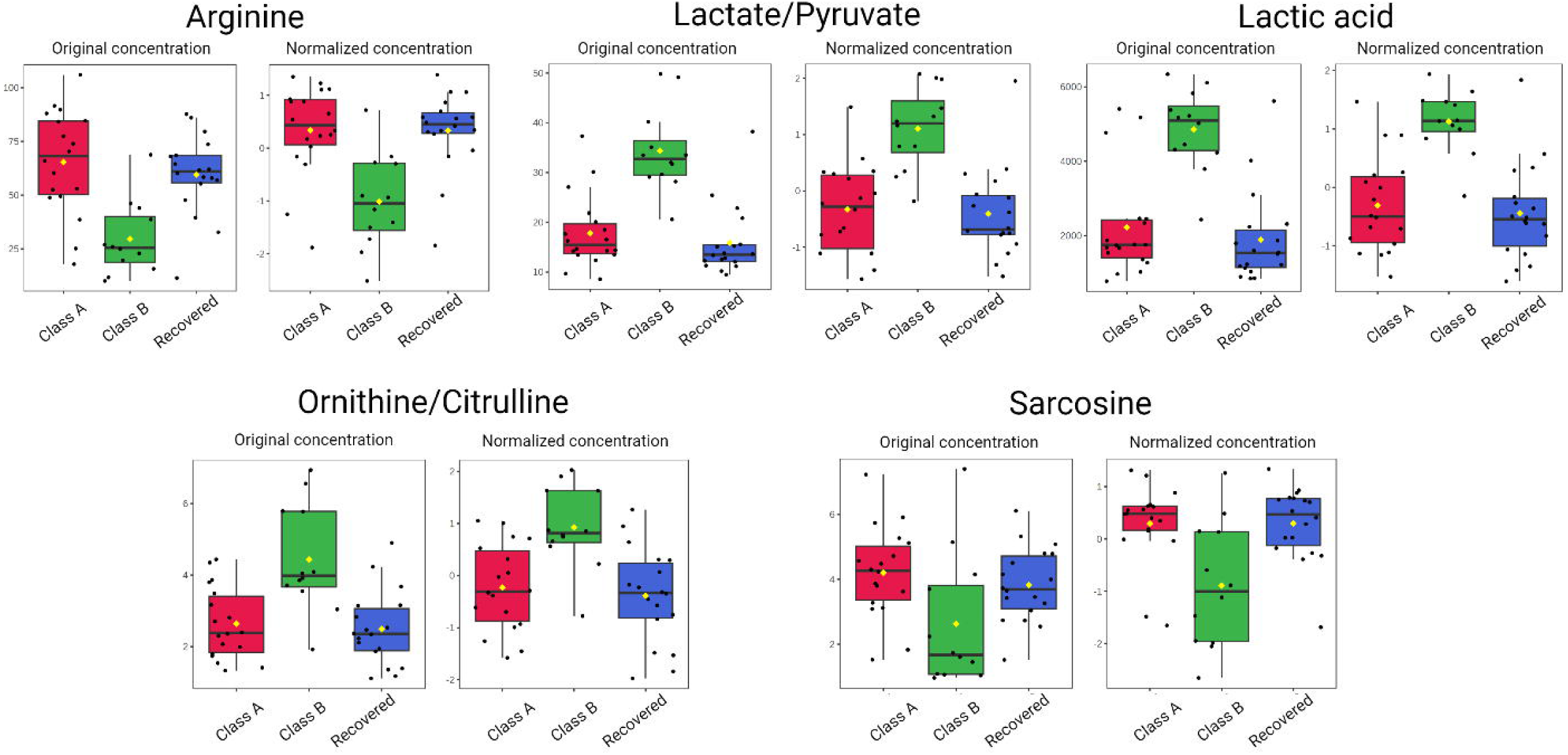
Box plots for some significantly altered metabolites (p < 0.05) in plasma of class A patients (less than five symptoms), class B patients (more than five symptoms), and recovered patients. The bar plots show the original and normalized values (mean +/-one standard deviation). Medians are indicated by horizontal lines within each box.

For differentiating class B long COVID patients from all other post-COVID-19 patients, the lactate/pyruvate ratio had the best performance (AUC: 0.95 (0.92 -0.97), sensitivity: 0.92 (0.87-0.97), specificity: 0.94 (0.91-0.98)), followed by the combination of the ornithine/citrulline ratio and uric acid (AUC: 0.92 (0.89-0.95), sensitivity: 0.83 (0.77-0.90), specificity: 0.84 (0.79-0.90)).

Class B patients reported symptoms falling in all the categories, while class A patients reported mainly neuropsychiatric symptoms. We also wanted to know whether the classification based on the group of symptoms would be most appropriate rather than the classification based on the number of symptoms. 18 patients had systemic symptoms, while 13 patients reported mostly neuropsychiatric associated disorders. Lactic acid, arginine, the lactate/pyruvate ratio, the ornithine/citrulline ratio and sarcosine were common in both types of long COVID classifications. Long saturated or monounsaturated LysoPCs were found to be increased in patients with neuropsychiatric disorders **(Supplementary Figure 2)**.

### Pathway analysis

Our pathway enrichment analysis **(Figure 5)** shows that the top five metabolic pathways significantly dysregulated (FDR<0.05) in post-COVID patients (relative to controls) were: phospholipids biosynthesis, gluconeogenesis, the glucose-alanine cycle, the Warburg effect, and taurine and hypotaurine metabolism. When comparing class B patients with those recovered, the top five metabolic pathways (FDR<0.05) were: pyruvate metabolism, gluconeogenesis, glycine and serine metabolism, urea cycle metabolism, and the Warburg effect.

**Figure 5:**
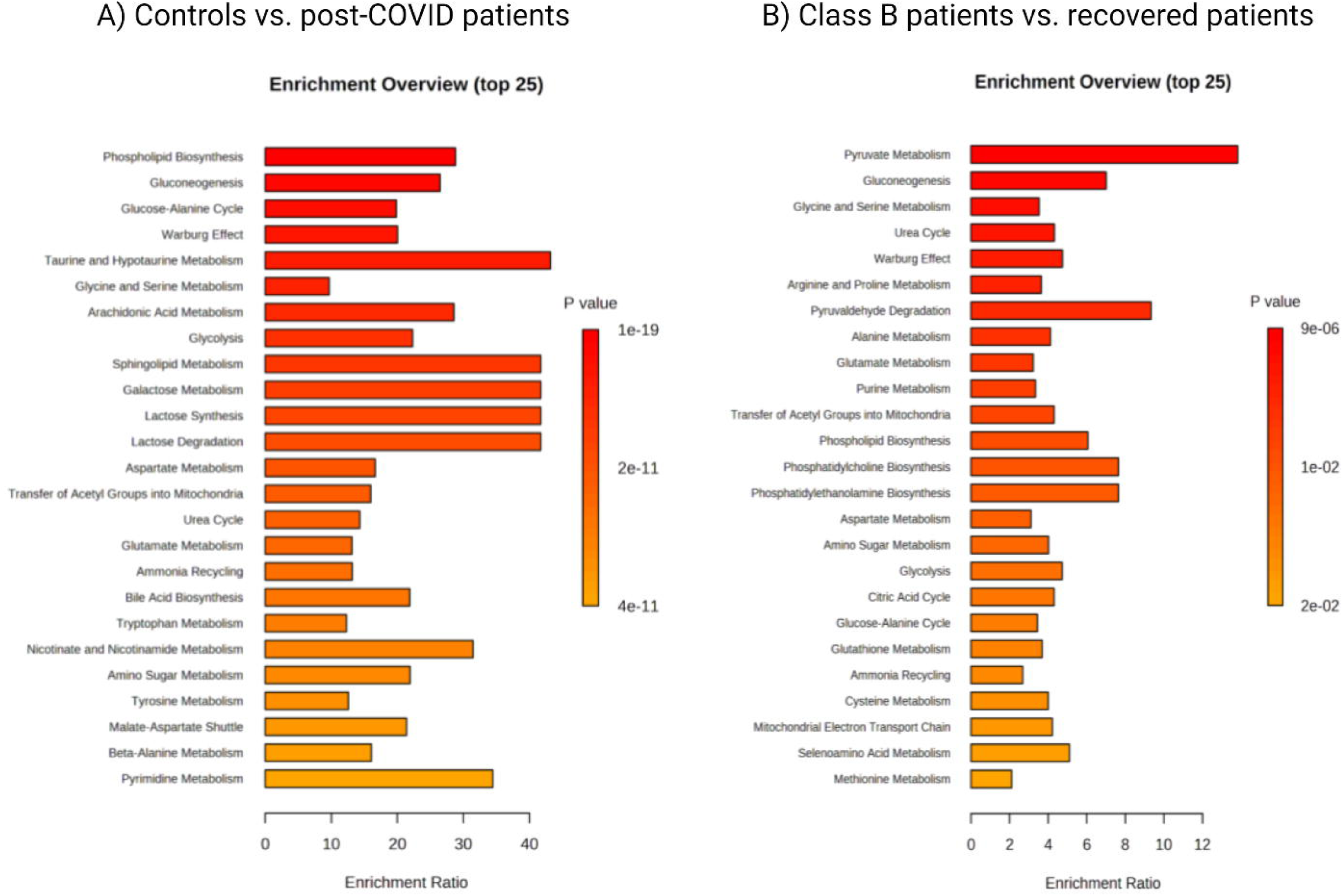
Metabolic pathway analysis. Predicted metabolic pathways with p-value_≤_0.05 are listed. A) controls vs. post-COVID-19 patients. B) class B patients vs. recovered patients.

### Plasma IL-17 and leptin

**Figure 6** shows plasma concentrations of IL-17 and leptin as measured by ELISA. IL-17 was significantly increased in class B patients relative to class A patients (Mann-Whitney test, p = 0.0073) and recovered patients (Mann-Whitney test, p = 0.002). Leptin did not show any statistically significant differences in the three-group comparison.

**Figure 6:**
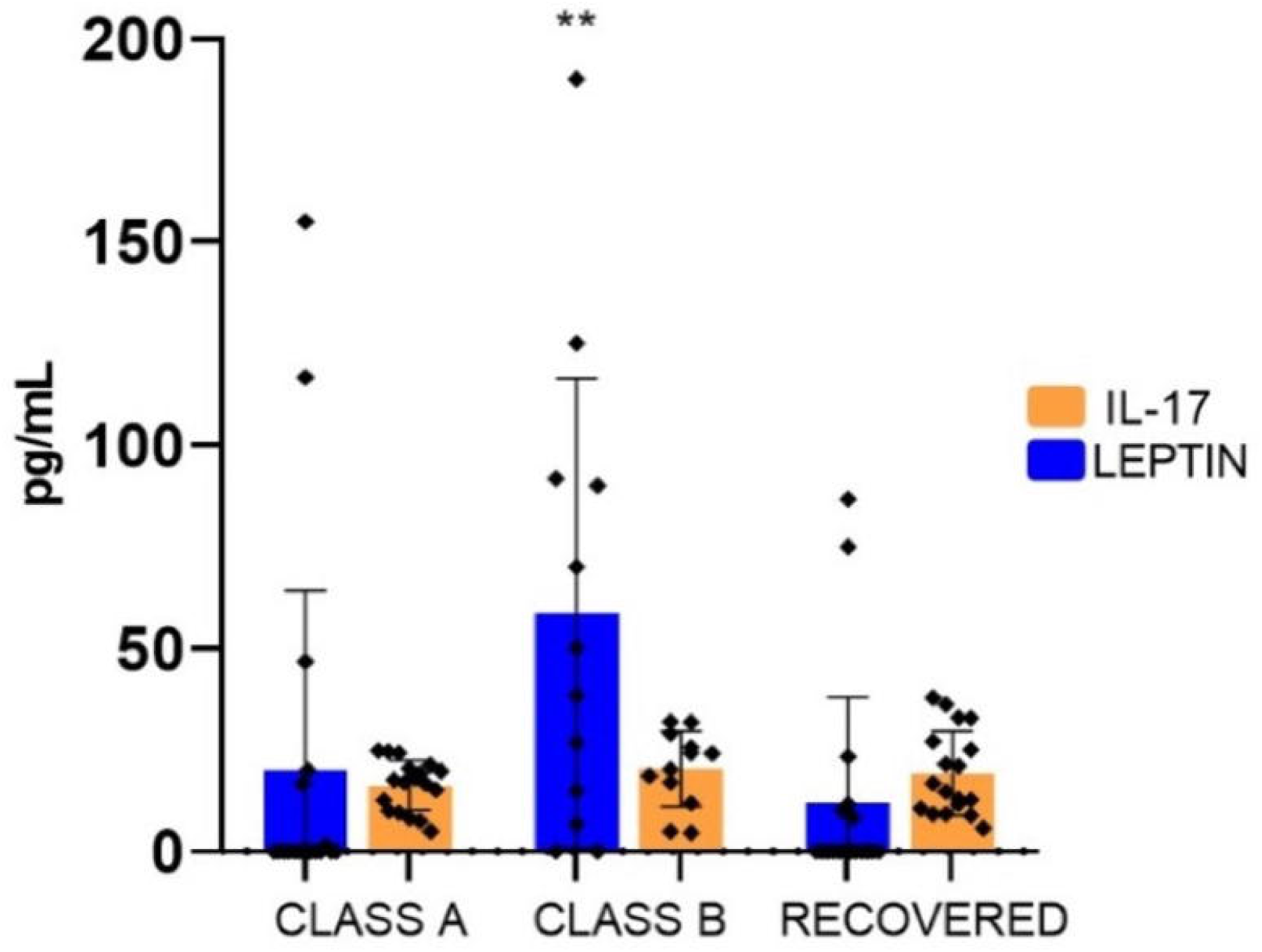
Concentrations of IL-17 and leptin measured by ELISA in post-COVID-19 patients. Graphs were constructed in GraphPad Prism v8.0. The ** p value <0.01 was calculated using Kruskall-Wallis tests with Dunńs post-tests.

## Discussion

Cumulative evidence from the last three years supports the dysregulation of metabolic and immune markers due to SARS-CoV-2 infection [20]. A retrospective cohort study has demonstrated that COVID-19 patients have a significantly higher risk to develop subsequent autoimmune diseases such as rheumatoid arthritis, ankylosing spondylitis, systemic sclerosis, type I diabetes mellitus, among others [21]. In the present work, our aim was to evaluate the persistence of long-term metabolic alterations in post-COVID-19 patients, as well as to measure immune markers that, when chronically produced, can trigger autoimmune diseases.

Since well-defined classification or diagnostic criteria are not available for the long COVID assessment, there is an urgent need for molecular methods able to stratify patients according to the severity of the symptoms they are experiencing. Quantitative and validated scales, such as HAM-A, HAM-D, MoCA and mMRC are considered gold standards for neurocognitive impairment and for dyspnea assessment. However, their practical utility could be limited for complex conditions such as long COVID where a broader range of self-reported symptoms with different severity and duration are present. It has been reported that some post-COVID-19 patients complain about extreme cognitive disorders (self-reported symptoms) but without any objective alterations, while others do not report symptoms but exhibit severe cognitive disorders after six to nine months following SARS-CoV-2 infection [22]. In our study, several symptoms were corroborated through objective measures, but with lower rates when using validated scales. Therefore, molecular markers are urgently needed for the correct classification of patients.

Our results revealed that 50% of analyzed plasma metabolites showed statistical differences between COVID-19 and post-COVID-19 phases. One of the most dysregulated metabolites was glucose. Montefusco *et al*. [23] reported glycemic abnormalities in recovered patients two months after the onset of disease. The hyperglycemic state has been reported to be even worse in hospitalized patients, pointing to a possible causal role of administered drug regimens, including remdesivir and corticosteroids. These drugs stimulate hepatic gluconeogenesis from amino acids released from muscles, which then inhibits glucose uptake [24].

A number of other metabolites were also found to be dysregulated. Increased plasma pyruvate levels could be both a consequence of glycolytic dysregulation and protein degradation. The increase in putrescine levels in the post-COVID phase may be an indicator of increased protein degradation to help fuel pyruvate metabolism.

Taurine and spermidine were found significantly decreased in the post-COVID phase, although a trend towards normalization was observed when compared with controls. Decreased levels of serum taurine have been observed in patients with Myalgic Encephalomyelitis/Chronic Fatigue Syndrome (ME/CFS) [25]. The depleted levels observed in post-COVID-19 phase could explain at least in part the fatigue, since taurine has multiple roles in skeletal muscle, the central nervous system, and energy metabolism. Nevertheless, based on our results, we did not find any correlation between fatigue or myalgia and taurine concentrations in post-COVID-19 patients. Holmes *et al*. [26] found that taurine levels were increased in post-COVID-19 patients, suggesting hepatic injury, hepatotoxicity, or muscle damage. However, the cohort evaluated in the Holmes study had a three-month follow-up after the initial infection, which is much shorter than the follow-up used in this study.

Furthermore, we observed increased levels of kynurenine (similar to levels in the COVID-19 acute phase), and a trend towards normalization in tryptophan and the kynurenine/tryptophan ratio in post-COVID-19 patients. This indicates that, although lower in magnitude, the inflammatory conditions attributable to the hyperactivation of this metabolic pathway are still present and may account for some persistent physiological symptoms in these patients.

The increase in glutamine (and decrease in glutamate levels) indicates a partial reestablishment of critical processes that took place during the COVID-19 infection phase, such as severe immunometabolic dysregulation. It is well known that SARS-CoV-2 induces metabolic reprogramming in host cells, similar to the Warburg effect in cancer, and a depletion of glutamine has been associated with its consumption for feeding TCA, and as a nitrogen source for nucleotide (ATP) synthesis [27]. On the other hand, glutamate is the most abundant neurotransmitter in the brain. A disruption within the glutamatergic pathway can lead to important neurological consequences, such as cognitive deficits [28]. The glutaminergic dysfunction could be associated with some psychiatric and neurologic symptoms like those reported in the present work.

Alterations in lipid metabolism are evident in most post-COVID-19 patients. These patients exhibited significantly higher levels of carnitine and some short, medium, and long acylcarnitines. These alterations have been largely associated with altered fatty acid metabolism, dysfunctional mitochondria-dependent lipid catabolism, and immune processes or the lysis of white blood cells. Similar results have been reported by Guntur *et al*. [29], pointing to mitochondrial dysfunction, as was also recognized during COVID-19 acute phase. Besides, decreased levels of LysoPC 16:0, LysoPC 17:0, LysoPC 18:0, and LysoPC 20:4, were found with respect to negative controls. These reductions have been reported in other inflammatory conditions [30] and other septic processes [31]. Depleted levels of lysophosphatidylcholines and phospholipid ethers, as well as depleted levels of PCs, can impede mitochondrial respiration, as has been also demonstrated in ME/CFS [32]. In line with the lipid dysregulation demonstrated by the targeted metabolomic analysis, routine clinical laboratory tests exhibited elevated levels of total cholesterol, triglycerides, and VLDL, as well as normal levels of HDL and LDL. Xu *et al*. [33] found increased LDL, triglycerides, total cholesterol, and decreased HDL in survivors of COVID-19, based on a large observational study with participants from the US Department of Veterans Affairs database compared to controls who had never tested positive for COVID-19.

As positive findings for the metabolic state of post COVID patients, we found that 30 metabolites fell within normal levels. Phenylalanine, which has been widely associated with sepsis and COVID disease severity [20] decreased to normal levels. Beta-hydroxybutyric acid and citric acid were also normalized, indicating partial recovery of the tricarboxylic acid cycle [20, 34]. Butyric acid and propionic acid, two short-chain fatty acids that were found to be altered during COVID-19 phase, also fell within normal levels in post-COVID-19 patients, probably indicating that the leaky gut phenomenon and gut dysbiosis detected during COVID infection could be partially reestablished [35]. Spermidine was also normalized. The decrease in spermidine levels could reflect a trend for normalization in overall redox balance. Although excessive levels of spermidine (as those reported in COVID-19 patients) trigger the production of superoxide radicals, optimal concentrations mitigate oxidative stress and diminish overall ROS production [36].

In addition, sphingomyelins and long-chain monounsaturated and saturated LysoPCs were found to be within normal levels. We previously noted altered sphingolipids levels during COVID-19 infection [20]. Sphingolipids play a crucial role in the regulation of signal transduction pathways and in certain pathological conditions, such as inflammation-associated illnesses and innate immune response.

In a recent report, Holmes *et al*. [26] found a high degree of interindividual variability in follow-up patients, reflecting the heterogeneity of post-COVID-19 patients and the fact that long COVID is a spectrum of disorders. Indeed, computationally modeling of the long COVID phenotype data based on electronic healthcare records found six distinct clusters, each with distinct profiles of phenotypic abnormalities [37]. Since symptom classification is still highly subjective, we decided to arbitrarily classify long COVID patients as: class A (less than five symptoms, mainly neuropsychiatric disorders), and class B (more than five symptoms, with a broad spectrum of systemic disorders).

We believe that metabolic information may complement, and partially explain the phenotypic differences among post-COVID-19 patients, and especially in long COVID patients. Xu *et al*. [38] classified recovered patients based on abnormal pulmonary functions, finding increased levels of triacylglycerols, phosphatidylcholines, prostaglandin E2, arginine, and decreased levels of betaine and adenosine in patients with abnormal pulmonary function.

In our work, lactic acid levels were increased in patients with more than five symptoms and systemic disorders (class B patients). Ghali *et al.* [39] found that patients with ME/CFS exhibited elevated blood lactate at rest. Mitochondrial dysfunction, with increased blood lactate, low levels of ATP, and increased levels of oxidative stress markers have been associated to these alterations [40], as well as relative deficiency of mitochondria type I fibers on muscle biopsies, and low intracellular pH during recovery phase [41–43]. De Boer *et al*. [44] also reported altered lactate levels in long COVID patients, suggesting that long COVID patients have significant impairment in fat beta-oxidation and increased blood lactate accumulation even during low-intensity exercise. In contrast, Guntur *et al*. [29] reported low levels of lactic acid and pyruvate in long COVID patients. However, this study was conducted in non-hospitalized patients who had recovered from COVID-19 in March 2020.

Increased level of the lactate/pyruvate ratio in class B patients is another important indicator of mitochondrial dysfunction. The lactate/pyruvate ratio has been proposed as a marker for mitochondrial disorders since it indirectly reflects the NADH/NAD+ redox state[45], lipid metabolism (fat oxidation), and ATP generation. In our study, both markers (lactate and the lactate/pyruvate ratio) were found positively correlated with fatigue, myalgia and arthralgias (Spearman correlation, R> 0.6, p< 0.05) **(Supplementary Figure 3).**

The increased ornithine/citrulline ratio level in class B patients reflects abnormal metabolic activity in the urea cycle. It is notable that Yamano *et al*. [46] reported a similarly increased ornithine/citrulline ratio in CFS patients. An adequate balance of citrulline and ornithine is vital for the clearance of ammonia via urea cycle [47]. If ammonia accumulates intracellularly, the aerobic utilization of pyruvate to feed the TCA cycle is inhibited, resulting in lactate production, which further contributes to fatigue.

In addition, class B patients had decreased levels of arginine. The reduced bioavailability of arginine to produce adequate levels of NO in endothelial cells and vascular tissues leads to the impairment of multiple physiological functions of skeletal muscles, including contractile functions, and muscle repair. This decreased level is not a residual effect of COVID-19, as the paired study (COVID-19/post-COVID-19) showed normal arginine levels. Arginine is also a substrate for ornithine production by arginase. It is well known that under certain inflammatory conditions, arginase activity is increased [48], producing an excess of ornithine and an imbalance in the urea cycle.

Sarcosine was found decreased in class B patients. Previously, Fraser *et al*. [49] found that sarcosine was depressed in COVID-19 patients. Sarcosine plays a vital role in immune functions, as it activates autophagy and the removal of damaged cells. A reduced amount of plasma sarcosine could in part lead to a sustained inflammatory process.

Previous studies have pointed to the persistent immune dysregulation following COVID-19 infection [50]. We found increased levels of monocytes in class B patients. Nuber-Champier *et al.* [51]found that monocyte percentage in the acute phase of the disease allowed them to distinguish between patients with anosognosia for memory deficits in the chronic phase (6–9 months after SARS-CoV-2 infection) and nosognosic patients.

We also measured IL-17 levels in post-COVID-19 patients since it is well known that this cytokine is persistently altered in several chronic inflammatory and autoimmune diseases [52], and previous reports have indicated an increased risk of such diseases in COVID-19 patients [21]. IL-17 is a proinflammatory cytokine mainly produced by T helper type 17 cells, playing a vital role in the regulation of host immune response against SARS-CoV-2. IL-17-induced dysregulated immune responses have been shown to potentially cause hyperinflammatory COVID-19 disease [53]. It has been reported that IL-17 downregulates protein phosphatase 6, resulting in increased arginase-1 expression in psoriatic keratinocytes [54]. IL-17A has been found to be associated with neurological sequelae and pulmonary fibrosis in post-COVID-19 patients [55, 56]. Fluctuations in IL-17 have been associated with fatigue and fatigue severity in ME/CFS patients [57].

Metabolomics is not only useful in providing a snapshot of transient physiological or pathophysiological processes taking place in a living organism, but it has also proven to be a powerful tool for proposing and monitoring therapeutic interventions. In the case of long COVID, a common situation worldwide is that patients have reported an absence of adequate support and a poor recognition of their condition, initially attributed to psychiatric issues. People with long COVID have tried a vast range of self-prescribed medicines, supplements, remedies, and dietary changes to manage the disease and to overcome the effects it has on their quality of life and work capacity. Based on our findings, some interventions could be tested for treating long COVID patients: 1) supplementation of taurine (reducing musculoskeletal disorders); 2) supplementation of arginine and/or citrulline (enhancing ammonia clearance and reducing blood lactate, as well as increasing arginine bioavailability for adequate NO production); 3) supplementation of glutamine (primary source for neurotransmitters and immune function balancing); 4) supplementation of antioxidants such as N-acetylcysteine or NAD+ (redox balance). Similarities found in our results with the ME/CFS pathophysiology may pave the way to common therapeutic interventions for both diseases.

We need to acknowledge several limitations with this study. The small sample size was due to the limited number of patients who agreed to participate. While several objective measures of mood and cognition (HAM-A, HAM-D, MoCA mMRC) were used, the sample size did not allow for stratification of patients according to the different test scores obtained, and only self-reported symptoms were used for sub-group classification. Furthermore, we were unable to have a detailed tracking of treatments, medications or alternative therapies during the period evaluated. This limited our interpretation with regard to the impact of pharmacological interventions on the metabolome. Moreover, patients from two different hospitals participated: one private hospital located in Chihuahua city, and one public hospital located in Zacatecas city. In general, private hospital patients have relatively high incomes while public hospital patients have lower incomes. Logistic regression models showed no effects of sex, age, comorbidities, vaccination status or severity during the acute phase in the metabolomic profile associated with long COVID. However, patients from the public hospital reported more systemic symptoms in general, while patients from the private hospital reported principally neuropsychiatric symptoms. A recent study found that patients diagnosed with a post-COVID-19 condition were more likely to be unemployed or on public health insurance, illustrating racial and social disparities in access to and experience with healthcare, at least in the USA [58]. Whether the socioeconomic conditions and lifestyles, along with causes of biological origin influence the metabolic phenoreversion of patients recruited in our study, needs to be further investigated. This is particularly important in countries with significant health system disparities and significant differences in population life conditions.

## Conclusions

To our knowledge, this study is the first describing quantitative metabolic perturbations two years after the initial acute COVID-19 infection using targeted metabolomics. The evolution of post-COVID-19 patients is different, and symptoms are associated to distinctive metabolic patterns resembling, to some extent, the ME/CSF condition. Moreover, the differences observed between the phenotypes of post-COVID-19 patients reveals potential biomarkers that could enable a more accurate and precise molecular classification of long COVID patients beyond classification via self-reported symptoms.

### List of abbreviations

COVID-19: Coronoravirus Disease 2019
LC-MS: Liquid Chromatography-mass spectrometry
FIA: Flow Injection Analysis
LysoPCs: Lysophosphatidylcholines
PACS: Post-Acute COVID-19 Syndrome
ME/CFS: Myalgic Encephalomyelitis/Chronic Fatigue Syndrome
IL-17: Interleukin 17
PC: Phosphatidylcholine
SM: Sphingomyelin
TCA: Tricarboxylic acid
HAM-A: Hamilton Anxiety Rating Scale
HAM-D: Hamilton Depression scale
MoCA: The Montreal Cognitive Assessment
mMRC: Medical Research Council

## Availability of data and materials

The datasets generated and analyzed during the current study are available in the Mendeley repository (https://data.mendeley.com) (doi: 10.17632/8zfdjsypd8.1).

## Ethics approval and consent to participate

The study was revised and approved by the Research and Ethics Committees of the Instituto Mexicano de Seguridad Social, with the registration number R-2022-3301-038, and Christus Muguerza del Parque Hospital (HCMP-CEI-15042020-3, and HCMP-CEI-28022022-A01).

## Consent for publication

Not required.

## Funding

CONACyT grant number 311880 “Identificación y validación de marcadores inmuno-metábolicos que incrementan la suscpetibilidad a desarrollar formas graves de infección por SARS-CoV-2”.

CONACyT grant number 319503 “Tamizaje masivo de pequeñas moléculas para la identificación de biomarcadores y candidatos terapéuticos con alto potencial de efectividad contra nuevas variantes de SARS-CoV-2 circulantes a nivel mundial”.

Christus Muguerza del Parque Hospital, Chihuahua, Mexico.

Instituto Mexicano de Seguridad Social.

Genome Alberta (a division of Genome Canada) grant number TMIC MC4, the Canadian Institutes of Health Research (CIHR) grant number FS 148461, the Canada Foundation for Innovation (CFI) grant number MSIF 35456.

## Authors’ contributions

Study concept and design: YLH and JMA. JCB, DAGL recruited study subjects and collected samples. Data acquisition: JZ, JPS, RM and EMM. Statistical analysis: MB, RM. Data interpretation: YLH, MB, JMA, JAL, CTC, DSW. Drafting the manuscript: YLH. Manuscript editing: CTC. DSW supervised the whole research and revised the original manuscript. All the authors contributed to the discussion and gave approval to the final version of the manuscript.

## Acknowledgements

JPE-D is a postdoctoral researcher at the Instituto Nacional de Medicina Genómica and received a fellowship from CONACYT in the program “Estancias Posdoctorales por México en Apoyo por SARS CoV2 (COVID-19, 2166969)”.

