## Supplementary table 1 for "The plasma metabolome of long COVID-19 patients two years after infection"

**Table S1:** Significant metabolites (FDR <0.05) dysregulated in the post-COVID-19 phase respect COVID-19 phase (paired analysis).

|  | t.stat | p.value | FDR |
| --- | --- | --- | --- |
| Phenylalanine | -7.1823 | 4.69E-06 | 0.00051144 |
| LysoPC a C26:1 | -6.6973 | 1.02E-05 | 0.0005532 |
| Threonine | 6.3956 | 1.67E-05 | 0.00060553 |
| Taurine | -6.0134 | 3.18E-05 | 0.00069505 |
| Spermidine | -5.8856 | 3.97E-05 | 0.00069505 |
| SM C18:1 | -5.8577 | 4.16E-05 | 0.00069505 |
| LysoPC a C28:0 | -5.8056 | 4.56E-05 | 0.00069505 |
| C18:2 | 5.6465 | 6.03E-05 | 0.00069505 |
| LysoPC a C16:0 | -5.6255 | 6.26E-05 | 0.00069505 |
| Proline | 5.5725 | 6.87E-05 | 0.00069505 |
| GLN/GLU | 5.5612 | 7.01E-05 | 0.00069505 |
| Sarcosine | -5.185 | 0.00013832 | 0.0011475 |
| Glutamine | 5.1552 | 0.00014609 | 0.0011475 |
| SM C18:0 | -5.1239 | 0.00015475 | 0.0011475 |
| alpha-Ketoglutaric acid | -5.1047 | 0.00016031 | 0.0011475 |
| Propionic acid | -5.0778 | 0.00016844 | 0.0011475 |
| LysoPC a C18:2 | 5.0322 | 0.00018325 | 0.001175 |
| Hippuric acid | 4.9138 | 0.00022832 | 0.0013826 |
| SM(OH) C16:1 | -4.6189 | 0.00039788 | 0.0022826 |
| LysoPC a C20:4 | -4.5321 | 0.00046946 | 0.0025585 |
| PC aa C38:0 | -4.352 | 0.00066337 | 0.0034432 |
| Glutamic acid | -4.0765 | 0.001133 | 0.0056135 |
| LysoPC a C17:0 | -3.9348 | 0.0014959 | 0.006669 |
| Choline | 3.9252 | 0.0015243 | 0.006669 |
| SM C16:1 | -3.9234 | 0.0015296 | 0.006669 |
| LysoPC a C26:0 | -3.8786 | 0.0016706 | 0.0067828 |
| LysoPC a C28:1 | -3.8757 | 0.0016801 | 0.0067828 |
| SM C16:0 | -3.6282 | 0.0027408 | 0.01067 |
| C12 | -3.5078 | 0.0034805 | 0.012952 |
| PC aa C36:6 | 3.4958 | 0.0035648 | 0.012952 |
| LysoPC a C18:0 | -3.4075 | 0.0042488 | 0.014939 |
| Spermine | -3.3795 | 0.0044926 | 0.015303 |
| Histidine | 3.3325 | 0.0049328 | 0.016293 |
| trans-Hydroxyproline | 3.2924 | 0.0053427 | 0.017128 |
| SM(OH) C24:1 | -3.2347 | 0.0059921 | 0.018661 |
| SM(OH) C22:1 | -3.1667 | 0.0068605 | 0.020392 |
| SM(OH) C14:0 | -3.1622 | 0.0069222 | 0.020392 |
| SM(OH) C22:2 | -3.1235 | 0.0074752 | 0.021417 |
| beta-Hydroxybutyric acid | -3.111 | 0.0076629 | 0.021417 |
| Citric acid | 3.0642 | 0.0084102 | 0.022806 |
| Tryptophan | 3.0542 | 0.0085783 | 0.022806 |
| C2 | -3.0249 | 0.0090917 | 0.023595 |
| Methionine | 2.9403 | 0.01075 | 0.02725 |
| Asymmetric√Ç¬†dimethylarginine | -2.9004 | 0.011632 | 0.028816 |
| C4OH | -2.8793 | 0.012129 | 0.029378 |
| Citrulline | 2.7679 | 0.015103 | 0.035788 |
| lactate/pyruvate | -2.7288 | 0.016311 | 0.037425 |
| Kynurenine/Tryptophan | -2.7208 | 0.016568 | 0.037425 |
| Valine | -2.713 | 0.016824 | 0.037425 |
| ornithine/citrulline | -2.6995 | 0.017273 | 0.037654 |
| Methionine-sulfoxide | -2.5779 | 0.021898 | 0.046802 |
| Glycine | 2.5601 | 0.022667 | 0.047514 |
| PC aa C40:6 | -2.5478 | 0.023216 | 0.047747 |
