## Supplementary table 2 for "The plasma metabolome of long COVID-19 patients two years after infection"

**Table S2:** Significant metabolites (FDR <0.05) dysregulated in post-COVID-19 phase respect to negative controls.

|  |  | **t.stat** | **p.value** | **-log10(p)** | **FDR** |
| --- | --- | --- | --- | --- | --- |
| LysoPC a C16:0 | Down | 9.9959 | 1.3081E-13 | 12.883 | 1.4259E-11 |
| gln/glu ratio | Up | -6.3847 | 5.0975E-08 | 7.2926 | 2.7781E-06 |
| LysoPC a C18:0 | Down | 6.2502 | 8.3012E-08 | 7.0809 | 3.0161E-06 |
| C18:2 | Up | -5.9722 | 2.2677E-07 | 6.6444 | 6.1795E-06 |
| Sarcosine | Down | 5.7633 | 4.8061E-07 | 6.3182 | 1.0477E-05 |
| Glutamine | Up | -5.2443 | 3.0425E-06 | 5.5168 | 4.2777E-05 |
| Glucose | Up | -5.2416 | 3.071E-06 | 5.5127 | 4.2777E-05 |
| Taurine | Down | 5.2353 | 3.1396E-06 | 5.5031 | 4.2777E-05 |
| Choline | up | -5.1187 | 4.7275E-06 | 5.3254 | 5.7256E-05 |
| LysoPC a C17:0 | Down | 4.9023 | 1.0033E-05 | 4.9986 | 0.00010936 |
| Kynurenine | Up | -4.3141 | 7.3619E-05 | 4.133 | 0.0007295 |
| Glutamic acid | Down | 4.1351 | 0.00013253 | 3.8777 | 0.0012038 |
| LysoPC a C16:1 | Down | 4.0618 | 0.00016812 | 3.7744 | 0.0014096 |
| Pyruvic acid | Up | -3.9811 | 0.00021796 | 3.6616 | 0.001697 |
| LysoPC a C20:4 | Down | 3.8703 | 0.00031014 | 3.5084 | 0.0022537 |
| C18:1 | Up | -3.6428 | 0.0006311 | 3.1999 | 0.0042994 |
| Kynurenine/Tryptophan | Up | -3.4694 | 0.0010696 | 2.9708 | 0.0068583 |
| Threonine | Up | -3.4426 | 0.0011594 | 2.9358 | 0.0070205 |
| C10:2 | Up | -3.417 | 0.0012514 | 2.9026 | 0.0071791 |
| Putrescine | Up | -3.2269 | 0.0021884 | 2.6599 | 0.011453 |
| LysoPC a C14:0 | Down | 3.224 | 0.0022066 | 2.6563 | 0.011453 |
| PC aa C32:2 | Down | 3.0629 | 0.0034958 | 2.4565 | 0.01732 |
| PC aa C40:2 | Up | -2.9939 | 0.0042409 | 2.3725 | 0.020098 |
| C10:1 | up | -2.8181 | 0.0068584 | 2.1638 | 0.031148 |
| PC ae C36:0 | Up | -2.7186 | 0.008938 | 2.0488 | 0.03897 |
| C0 | up | -2.6778 | 0.0099471 | 2.0023 | 0.041701 |
| SM(OH) C22:1 | Down | 2.5756 | 0.01295 | 1.8877 | 0.049774 |
| lactate/pyruvate | Down | 2.5716 | 0.013081 | 1.8834 | 0.049774 |
| Ornithine | up | -2.5668 | 0.013243 | 1.878 | 0.049774 |
