## Supplementary Table 3 for "The plasma metabolome of long COVID-19 patients two years after infection"

**Table S3:** Logistic Regression Model - Summary of Each Feature and performance

|  | **Estimate** | **Std. Error** | **Z value** | **Pr(>\|z\|)** | **Odds** |
| --- | --- | --- | --- | --- | --- |
| (Intercept) | 1.13 | 0.62 | 1.824 | 0.068 | - |
| Myalgia | -1.724 | 0.602 | -2.866 | 0.004 | 0.18 |
| Ornithine/citrulline | -2.763 | 1.002 | -2.758 | 0.006 | 0.06 |
| Lactate/pyruvate | 3.076 | 0.969 | 3.176 | 0.001 | 21.66 |

|  | **AUC** | **Sensitivity** | **Specificity** |
| --- | --- | --- | --- |
| Training/Discovery | 0.95 (0.93-0.98) | 0.93 (0.91-0.96) | 0.94 (0.91-0.98) |
| 10-fold Cross-Validation | 0.94 (0.85-1.00) | 0.93 (0.93-1.00) | 0.95 (0.86-1.00) |
