## Supplementary figures and images for "The plasma metabolome of long COVID-19 patients two years after infection"

### Supplementary Figure 1

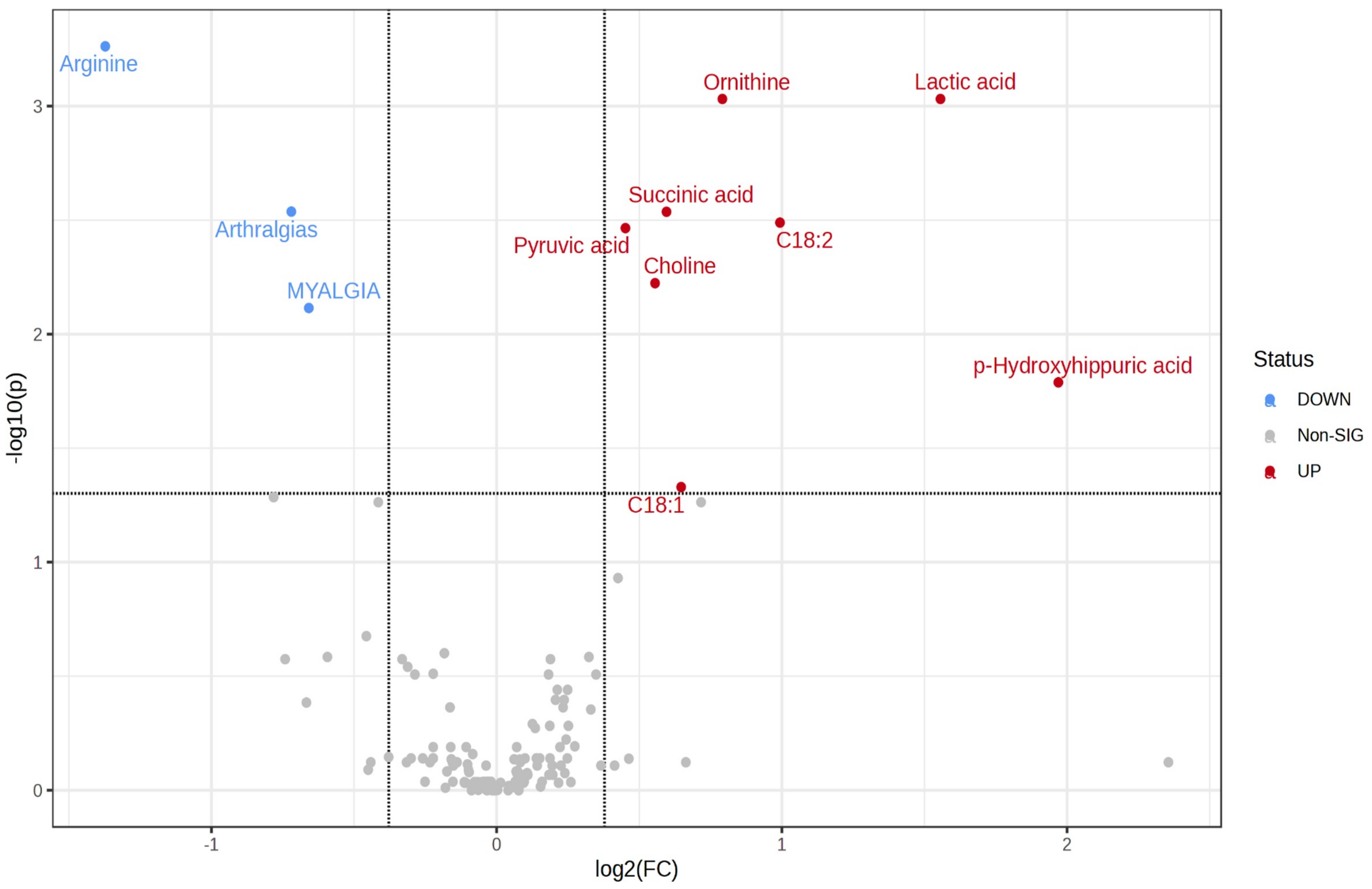

### Supplementary Figure 2

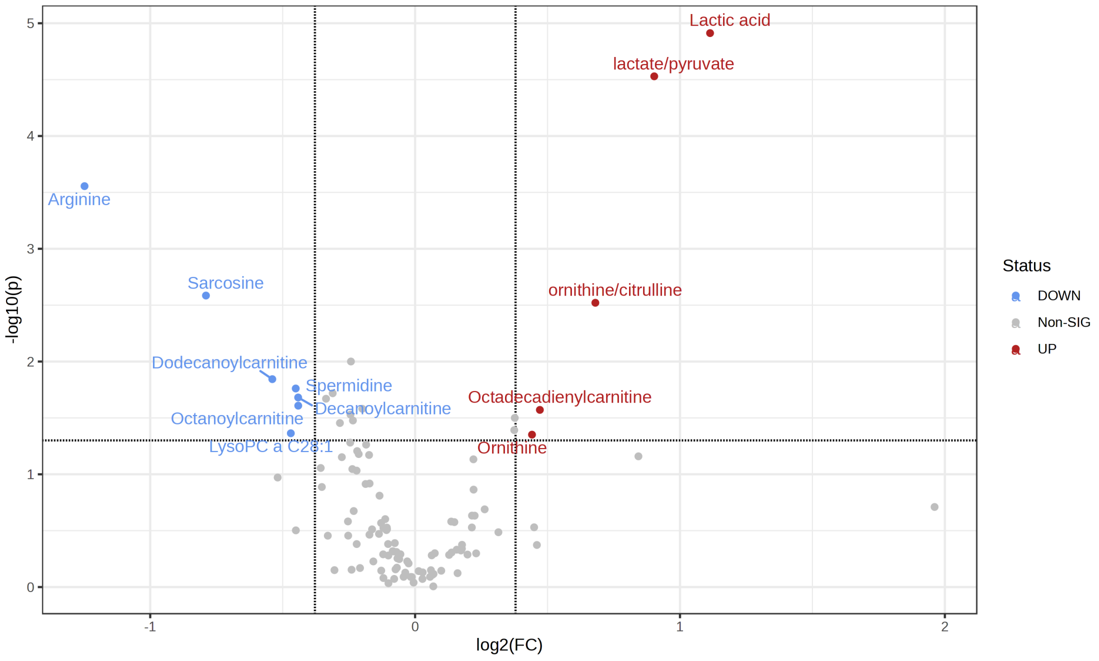

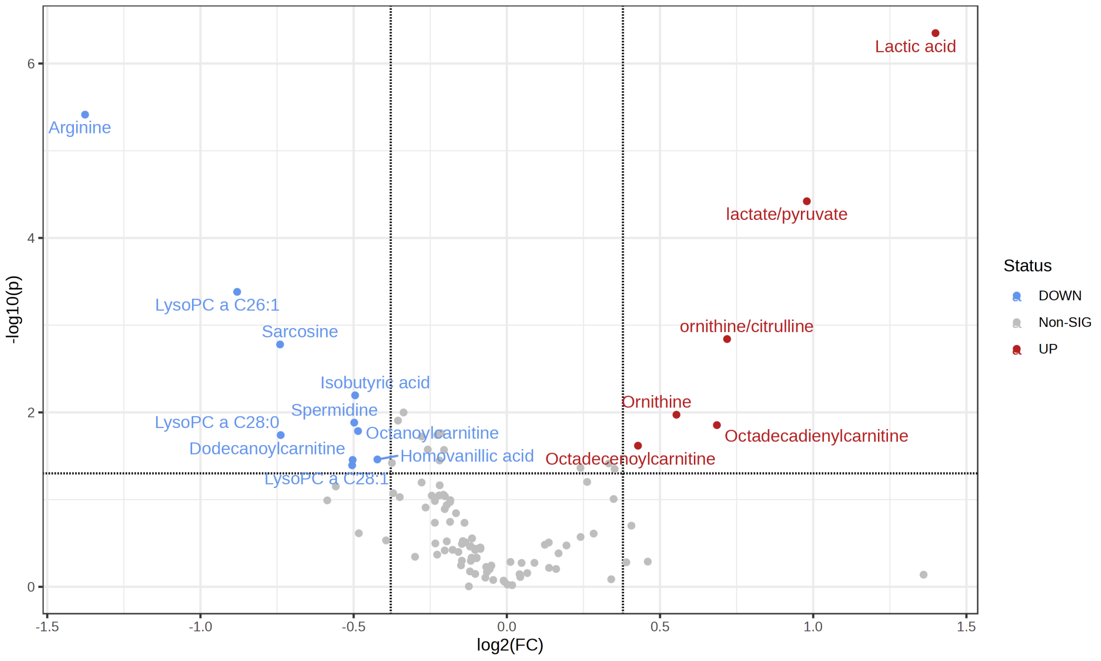

### Supplementary Figure 3

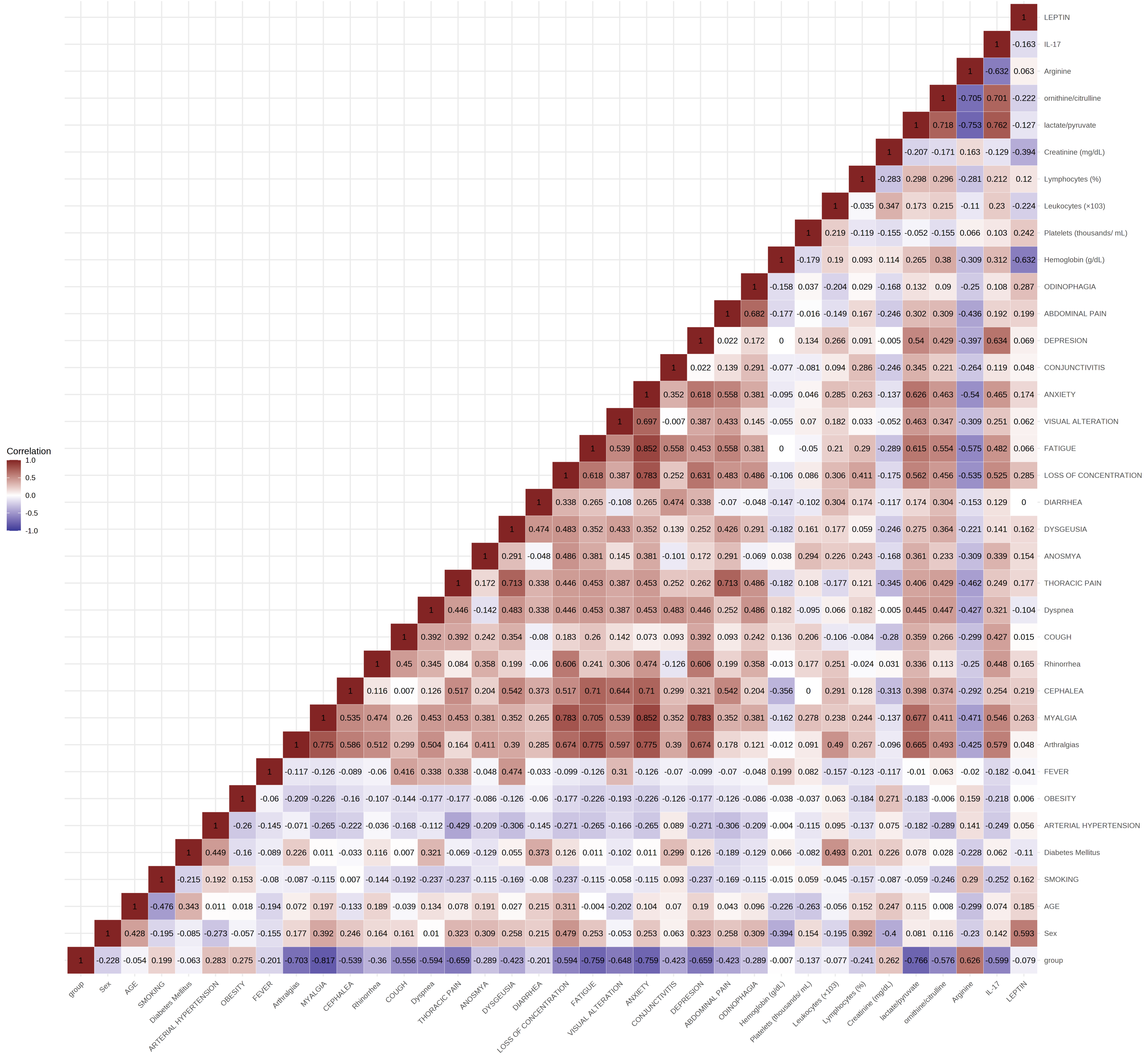
